# Classification of patients with osteoarthritis through clusters of comorbidities using 633,330 individuals from Spain

**DOI:** 10.1101/2022.09.22.22280234

**Authors:** Marta Pineda-Moncusí, Francesco Dernie, Andrea Dell’Isola, Anne Kamps, Jos Runhaar, Subhashisa Swain, Weiya Zhang, Martin Englund, Irene Pitsillidou, Victoria Y Strauss, Danielle E Robinson, Daniel Prieto-Alhambra, Sara Khalid

## Abstract

**Objectives:** To explore clustering of comorbidities among patients with a new diagnosis of osteoarthritis (OA) and estimate the 10-year mortality risk for each identified cluster.

**Methods:** This is a population-based cohort study of individuals with first incident diagnosis of OA of the hip, knee, ankle/foot, wrist/hand, or ‘unspecified’ site between 2006 and 2020, using SIDIAP (a primary care database representative from Catalonia, Spain). At the time of OA diagnosis, conditions associated with OA in the literature that were found in ≥1% of the individuals (n=35) were fitted into two cluster algorithms, K-means and latent class analysis (LCA). Models were assessed using a range of internal and external evaluation procedures. Mortality risk of the obtained clusters was assessed by survival analysis using Cox proportional hazards.

**Results:** We identified 633,330 patients with a diagnosis of OA. Our proposed best solution used LCA to identify four clusters: ‘Low-morbidity (relatively low number of comorbidities), ‘Back/neck pain plus mental health’, ‘Metabolic syndrome’ and ‘Multimorbidity’ (higher prevalence of all study comorbidities). Compared to the ‘Low-morbidity, the ‘Multimorbidity’ cluster had the highest risk of 10-year mortality (adjusted HR: 2.19 [95%CI: 2.15-2.23]), followed by ‘Metabolic syndrome’ (adjusted HR: 1.24 [95%CI: 1.22-1.27]]) and ‘Back/neck pain plus mental health’ (adjusted HR: 1.12 [95%CI: 1.09-1.15]).

**Conclusion:** Patients with a new diagnosis of OA can be clustered into groups based on their comorbidity profile, with significant differences in 10-year mortality risk. Further research is required to understand the interplay between OA and particular comorbidity groups, and the clinical significance of such results.

**Key Messages:**

- Patients with newly diagnosed osteoarthritis can by classified into different clusters by their comorbidity patterns.
- Such classification can help identify ‘high-risk’ patients who require more intense attention from healthcare providers.
- The main patient sub-groups were ‘Low-morbidity’, ‘Back/neck pain plus mental health’, ‘Metabolic syndrome’ and ‘Multimorbidity’.

## Introduction

Osteoarthritis (OA) is a common chronic condition affecting about 250 million people worldwide^1^. The progressive degenerative nature of the disease causes functional impairment, often severe pain, and loss of quality of life^2^.

Given its chronic nature, OA often coexists alongside other chronic conditions (a.k.a. comorbidities). A systematic review has shown that patients with OA are more likely to have multiple conditions compared to patients without OA^3^, and further studies have shown that this increased likelihood exists both in the years preceding a diagnosis of OA, as well as in the years after^4^.

The co-existence of two or more chronic conditions is termed multimorbidity^5^, and is estimated to affect between 19-27% of the UK general population^6-8^. Studies have shown that increasing multimorbidity is associated with lower socioeconomic status^7,8^ and increasing age^7^, and drives higher healthcare utilisation including primary care usage, prescription costs and hospitalisation^8,9^. There is a growing realisation that understanding multimorbidity is important, both in clinical practice and in the development of clinical guidelines^10,11^.

Within the context of multimorbidity, there is increasing recognition of the concept of comorbidities existing in groups or ‘clusters’^12^. Examining the exact conditions which co-exist within an individual, rather than simply the number of comorbidities, would allow us to understand whether a patient’s chronic comorbid conditions are ‘concordant’ (may be treated with a unified approach), or ‘discordant’ (may worsen or compete with treatments for individual conditions)^13^, with important repercussions for the treatment of that individual, including polypharmacy^14^.

Clustering of comorbidities among individuals with OA has only recently started to be explored. Studies examining general multimorbidity have shown that musculoskeletal problems including OA are very common among people with multimorbidity^15^, and often cluster with cardiovascular disease^16,17^. OA is a particularly common contributor to multimorbidity among the elderly^17^. Such multimorbidity involving OA not only leads to further negative effects on quality of life, but also complicate treatment and increase requirements for analgesia^18^. With respect to the clustering of comorbidities specifically in individuals with OA, one large scale study in the UK has recently demonstrated five distinct clusters of comorbidities which predicted general practice (GP) consultation rates and mortality^19^.

In this study we used machine learning techniques to examine large-scale data from patients with OA to further explore clustering of comorbidities in primary care patients with OA in the Spanish population.

## Methods

### Study design, setting and data sources

We conducted a population-based cohort study using the Information System for Research in Primary Care (SIDIAP) healthcare database, which collects de-identified patient records from 279 primary care providers in Catalonia, Spain, covering around 80% of the Catalan population, or 5.8 million people^20^. This study forms part of the Comorbidities in Osteoarthritis (ComOA) project, the protocol for which has been published previously^21^.

### Participants and study size

We included all participants aged ≥18 years with a least one physician-recorded diagnosis of OA of the hip, knee, ankle/foot, wrist/hand, or ‘unspecified’ site between 1st of January 2006 to 31st of June 2020, using ICD-10 codes (International Classification of Diseases 10th revision). The index date (date of their first incidence diagnosis of OA) was identified for each participant, and participants were followed from this date. Participants were excluded if they did not have at least one year of data recorded prior to their index date, or if they had a specific non-OA diagnosis (soft-tissue disorders, other bone/cartilage diseases) at the same joint in the 12 months prior to or after the index OA/joint pain date.

### Outcomes

The outcomes of interest were 1) clusters of comorbidities in people with OA and 2) risk of mortality in 10 years.

Mortality follow-up: individuals were followed from the date of OA diagnosis until the earliest: 1) date of death, or 2) date of transfer out of catchment area or end-date of data availability in SIDIAP.

### Variables

#### Comorbidities

A comprehensive initial list of 58 comorbidities was informed by a literature review and by expert opinion (Table 1). The extraction of comorbidities from individuals was performed at the time of OA diagnosis.

**Table 1.** Baseline characteristics.

| External Variables |  |
| --- | --- |
| Patients | N=633,330 |
| **Sex (n(%)):** |  |
| Female | 425,826 (67.2%) |
| Male | 207,504 (32.8%) |
| **Age (mean (sd))** | 67.3 (13.0) |
| **Body mass index (mean(sd))** | 29.3 (5.3) |
| NA | 541,318 |
| **Body mass index by categories (n(%)):** |  |
| <18.5 | 524 (0.57%) |
| 18.5 to 24.9 | 17791 (19.3%) |
| 25 to 29.9 | 36998 (40.2%) |
| 30+ | 36699 (39.9%) |
| NA | 541,318 |
| **QMEDEA deprivation index (n(%)):** |  |
| Urban area 1 (less deprived area) | 85,843 (13.6%) |
| Urban area 2 | 87,071 (13.8%) |
| Urban area 3 | 90,159 (14.2%) |
| Urban area 4 | 89,832 (14.2%) |
| Urban area 5 (more deprived area) | 82,812 (13.1%) |
| Unknown Urban area | 72,498 (11.5%) |
| Rural area | 124,629 (19.7%) |
| NA | 486 |
| **Smoke status (n(%)):** |  |
| Never smokers | 340834 (64.8%) |
| Current smokers | 79004 (15.0%) |
| Ex-smokers | 106546 (20.2%) |
| NA | 106,946 |
| **Risk of alcoholism (n(%)):** |  |
| None/low | 60794 (61.7%) |
| Moderate | 36523 (37.1%) |
| High/alcoholic | 1198 (1.2%) |
| NA | 534,815 |

#### Other variables

A set of external characteristics (i.e., not included in the cluster algorithms) from individuals at index-date was used to describe the obtained clusters: sex, age, body mass index (BMI), socioeconomic status, smoking and alcohol risk. BMI was classified into four categories: 1 (underweight, BMI<18.5); 2 (healthy weight, 18.5 ≤ BMI < 25); 3 (overweight, 25 ≤ BMI <30); 4 (obese, BMI ≥ 30). Socioeconomic status of the individuals was measured using of the MEDEA deprivation index^22^: urban areas are represented as quintiles (i.e., from U1 to U5), were U1 are the less deprived areas and U5 are the most deprived, and rural areas (R) are diferentiated^23^.

### Statistical methods

The external characteristics of participants and the prevalence of each comorbidity were described at the index date. Comorbidities found in less than 1% of the study population were excluded: their inclusion in the cluster algorithms increases the running times and the sample noise rather than drive to specific cluster solutions. Individuals were then classified into different clusters using K-means and latent class analysis (LCA) algorithms.

K-means is a type of ‘hard’ clustering approach, where individuals can only belong to one group in a binary fashion^24,25^. In order to identify the optimal number of clusters (*k*), we evaluated the clusters both internally, using within-cluster sum of squares (WCSS) and externally, by validating the clusters based on the external characteristics of the participants within each cluster: we selected the three cluster solutions from the WCSS before their change became lower than ±1 standard deviation (compared to the prior value); and then we explored them by assessing the prevalence of the comorbidities in each of the clusters and the external variables.

In contrast to ‘hard’ clustering approaches, ‘soft’ approaches such as LCA^26,27^ return the probability of an individual belonging to a particular group/cluster. To identify the potential optimal *k*, we compared the performance of the models from k=1 to k=10, using a number of metrics: entropy R^28,29^; goodness of fit tests ^30,31,32^; and log-likelihood ratio. Participants were assigned to the cluster with the higher posterior probability and then internally and externally validated using the same strategy as K-means, except for the initial selection of *k* clusters, that in this case depended on the lack of change (>±1 standard deviation) of entropy and goodness of fit tests and likelihood values.

For an easier understanding of the results, both K-means and LCA resulting clusters were assigned to a tag/identifier that clinically represents the grouped patients.

To calculate the 10-year mortality risk for each cluster, survival analysis^33^ was performed using Kaplan-Meier to plot the unadjusted curve of mortality in each cluster, and the Cox proportional hazards to calculate hazard ratios (HRs). We report the HRs with 95% confidence intervals (CI), both unadjusted and adjusted for age and sex.

All statistical analyses were conducted using R 4.1.1 for Windows.

## Results

A total of 633,330 patients were identified with a diagnosis of OA between 1st January 2006 and 31st June 2020. Our cohort was predominantly female (67.2%), with a mean age of 67.3 years. A large proportion of participants were either overweight (40.2%) or obese (39.9%). The baseline characteristics of the cohort is given in Table 1.

After exclusion of comorbidities with a prevalence of less than 1% (Table 2), a total of 35 comorbidities were included in the cluster analysis. The most common comorbidities were back/neck pain (33.6%) and hypertension (23.5%).

**Table 2.**
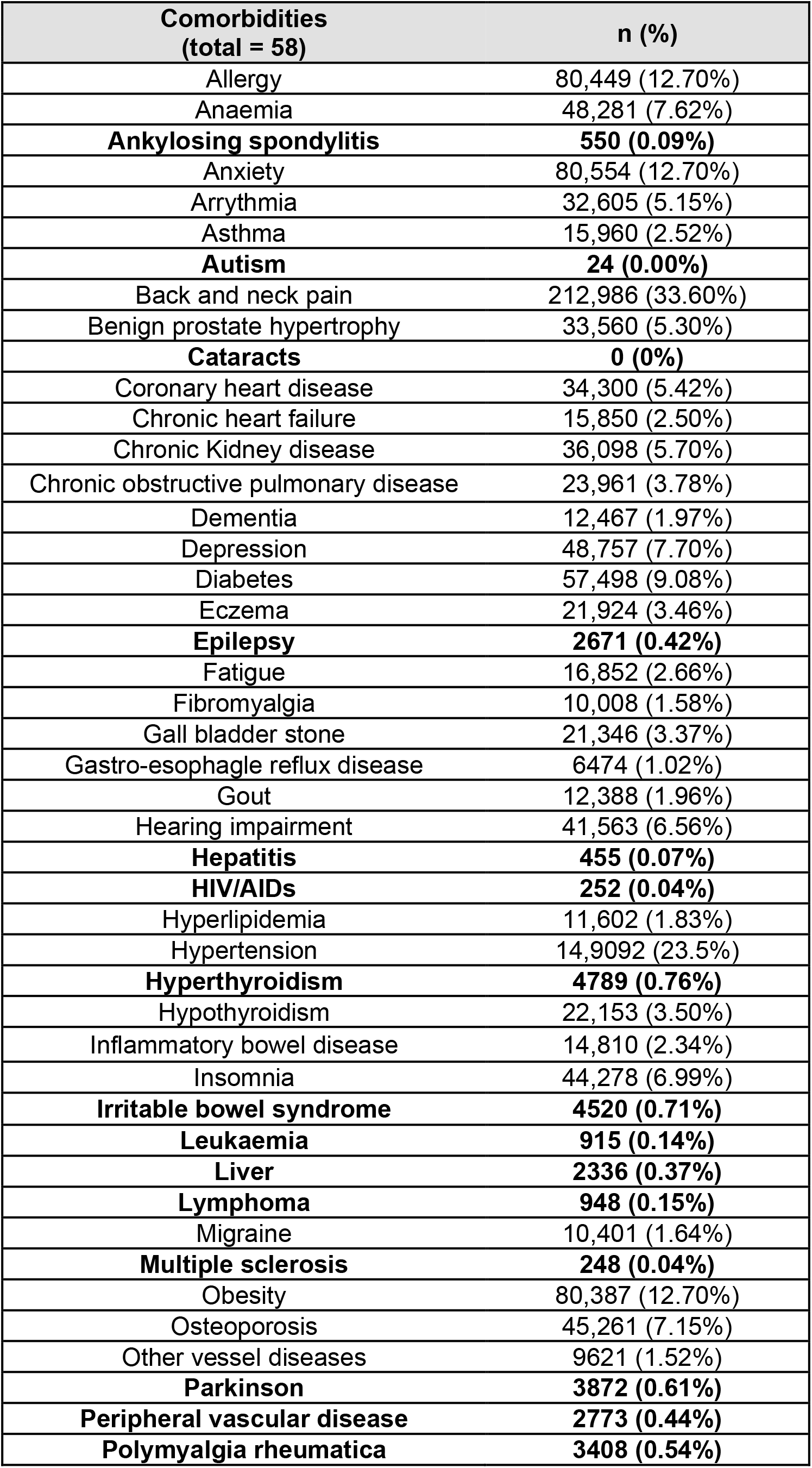
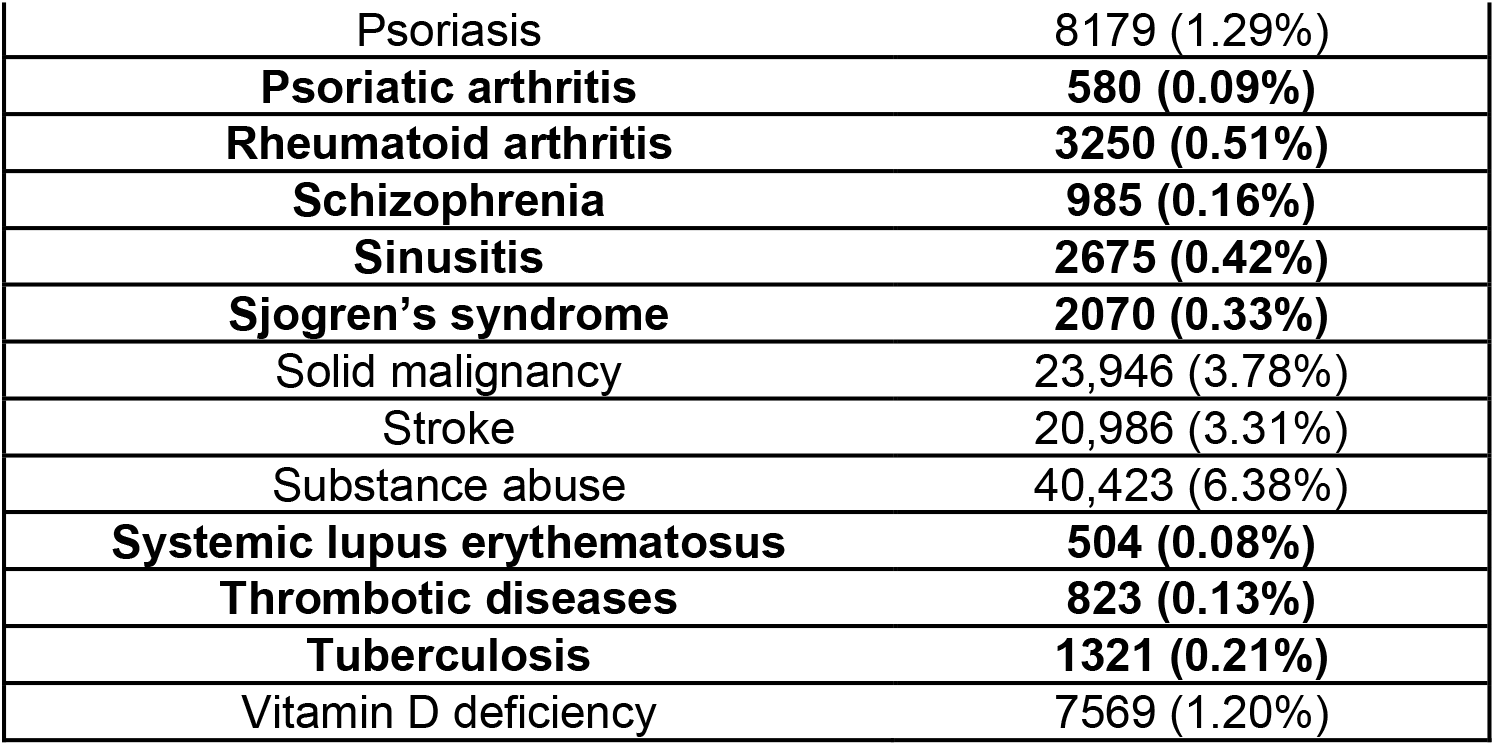
Prevalence of individual comorbidities at baseline. Comorbidities with a prevalence of <1%, excluded from final cluster analyses, are highlighted in bold.

| **Comorbidities**<br>**(total = 58)** | **n (%)** |
| --- | --- |
| Allergy | 80,449 (12.70%) |
| Anaemia | 48,281 (7.62%) |
| **Ankylosing spondylitis** | **550 (0.09%)** |
| Anxiety | 80,554 (12.70%) |
| Arrhythmia | 32,605 (5.15%) |
| Asthma | 15,960 (2.52%) |
| **Autism** | **24 (0.00%)** |
| Back and neck pain | 212,986 (33.60%) |
| Benign prostate hypertrophy | 33,560 (5.30%) |
| **Cataracts** | **0 (0%)** |
| Coronary heart disease | 34,300 (5.42%) |
| Chronic heart failure | 15,850 (2.50%) |
| Chronic Kidney disease | 36,098 (5.70%) |
| Chronic obstructive pulmonary disease | 23,961 (3.78%) |
| Dementia | 12,467 (1.97%) |
| Depression | 48,757 (7.70%) |
| Diabetes | 57,498 (9.08%) |
| Eczema | 21,924 (3.46%) |
| **Epilepsy** | **2671 (0.42%)** |
| Fatigue | 16,852 (2.66%) |
| Fibromyalgia | 10,008 (1.58%) |
| Gall bladder stone | 21,346 (3.37%) |
| Gastro-esophagel reflux disease | 6474 (1.02%) |
| Gout | 12,388 (1.96%) |
| Hearing impairment | 41,563 (6.56%) |
| **Hepatitis** | **455 (0.07%)** |
| **HIV/AIDs** | **252 (0.04%)** |
| Hyperlipidemia | 11,602 (1.83%) |
| Hypertension | 14,9092 (23.5%) |
| **Hyperthyroidism** | **4789 (0.76%)** |
| Hypothyroidism | 22,153 (3.50%) |
| Inflammatory bowel disease | 14,810 (2.34%) |
| Insomnia | 44,278 (6.99%) |
| **Irritable bowel syndrome** | **4520 (0.71%)** |
| **Leukaemia** | **915 (0.14%)** |
| **Liver** | **2336 (0.37%)** |
| **Lymphoma** | **948 (0.15%)** |
| Migraine | 10,401 (1.64%) |
| **Multiple sclerosis** | **248 (0.04%)** |
| Obesity | 80,387 (12.70%) |
| Osteoporosis | 45,261 (7.15%) |
| Other vessel diseases | 9621 (1.52%) |
| **Parkinson** | **3872 (0.61%)** |
| **Peripheral vascular disease** | **2773 (0.44%)** |
| **Polymyalgia rheumatica** | **3408 (0.54%)** |
| Psoriasis | 8179 (1.29%) |
| **Psoriatic arthritis** | **580 (0.09%)** |
| **Rheumatoid arthritis** | **3250 (0.51%)** |
| **Schizophrenia** | **985 (0.16%)** |
| **Sinusitis** | **2675 (0.42%)** |
| **Sjogren's syndrome** | **2070 (0.33%)** |
| Solid malignancy | 23,946 (3.78%) |
| Stroke | 20,986 (3.31%) |
| Substance abuse | 40,423 (6.38%) |
| **Systemic lupus erythematosus** | **504 (0.08%)** |
| **Thrombotic diseases** | **823 (0.13%)** |
| **Tuberculosis** | **1321 (0.21%)** |
| Vitamin D deficiency | 7569 (1.20%) |

### Clustering by K-means

Internal validation using WCSS showed us that the biggest reduction of the within clusters distance occur up to k=4, and solutions initially selected as potentially optimal were k=4, k=5 and k=6 (representative of the number of groups which participants could be clustered into, I.e., 4-cluster, 5-cluster and 6-cluster solutions, respectively) (Supplementary Figure S1A). However, no significant improvement was observed in 5- and 6-cluster solutions after assessing the distribution of comorbidity patterns within each cluster solution and the external variables. Thus, the 4-cluster solution was selected as the best K-means solution (Table 3).

**Table 3.** Survival analysis for 10-year mortality in 4-cluster solutions of a) K-means and B) Latent Class Analysis:

| Cluster number | Cluster name | Crude OR [95CI%] | Adjusted OR [95CI%] |
| --- | --- | --- | --- |
| 3 | Low-morbidity | Ref. | Ref. |
| 1 | Back and neck pain | 0.72 [0.70 - 0.73] | 0.93 [0.91 - 0.95] |
| 4 | Mental health | 0.87 [0.84 - 0.89] | 1.21 [1.18 - 1.24] |
| 2 | Metabolic Syndrome | 1.62 [1.60 - 1.65] | 1.18 [1.16 - 1.20] |
| - | Age | - | 1.14 [1.14 - 1.14] |
| - | Sex (male) | - | 1.73 [1.71 - 1.76] |

Abbreviations: CI, confidence intervals; Ref., Reference group; OR, Odds Ratio.

b) Latent Class Analysis
| Cluster number | Cluster name | Crude OR [95CI%] | Adjusted OR [95CI%] |
| --- | --- | --- | --- |
| 3 | Low-morbidity | Ref. | Ref. |
| 2 | 'Back and neck pain' plus 'mental health' | 0.85 [0.83 - 0.87] | 1.12 [1.09 - 1.15] |
| 4 | Metabolic Syndrome | 1.70 [1.67 - 1.74] | 1.24 [1.22 - 1.27] |
| 2 | Multimorbidity | 5.71 [5.61 - 5.81] | 2.19 [2.15 - 2.23] |
| - | Age | - | 1.13 [1.13 - 1.13] |
| - | Sex (male) | - | 1.68 [1.66 - 1.70] |

Abbreviations: CI, confidence intervals; Ref., Reference group; OR, Odds Ratio.

For k=4, distribution of comorbidity patterns led us to identify the following clusters (ordered from the largest to the lowest size): ‘low-morbidity’ (n= 302,733, 47.8%), ‘metabolic syndrome’ (n= 125,590,19.8%), ‘back and neck pain’ (n= 124,496, 19.7%), and ‘mental health’ (n= 80,511, 12.7%). (Figure 1A)

**Figure 1.**
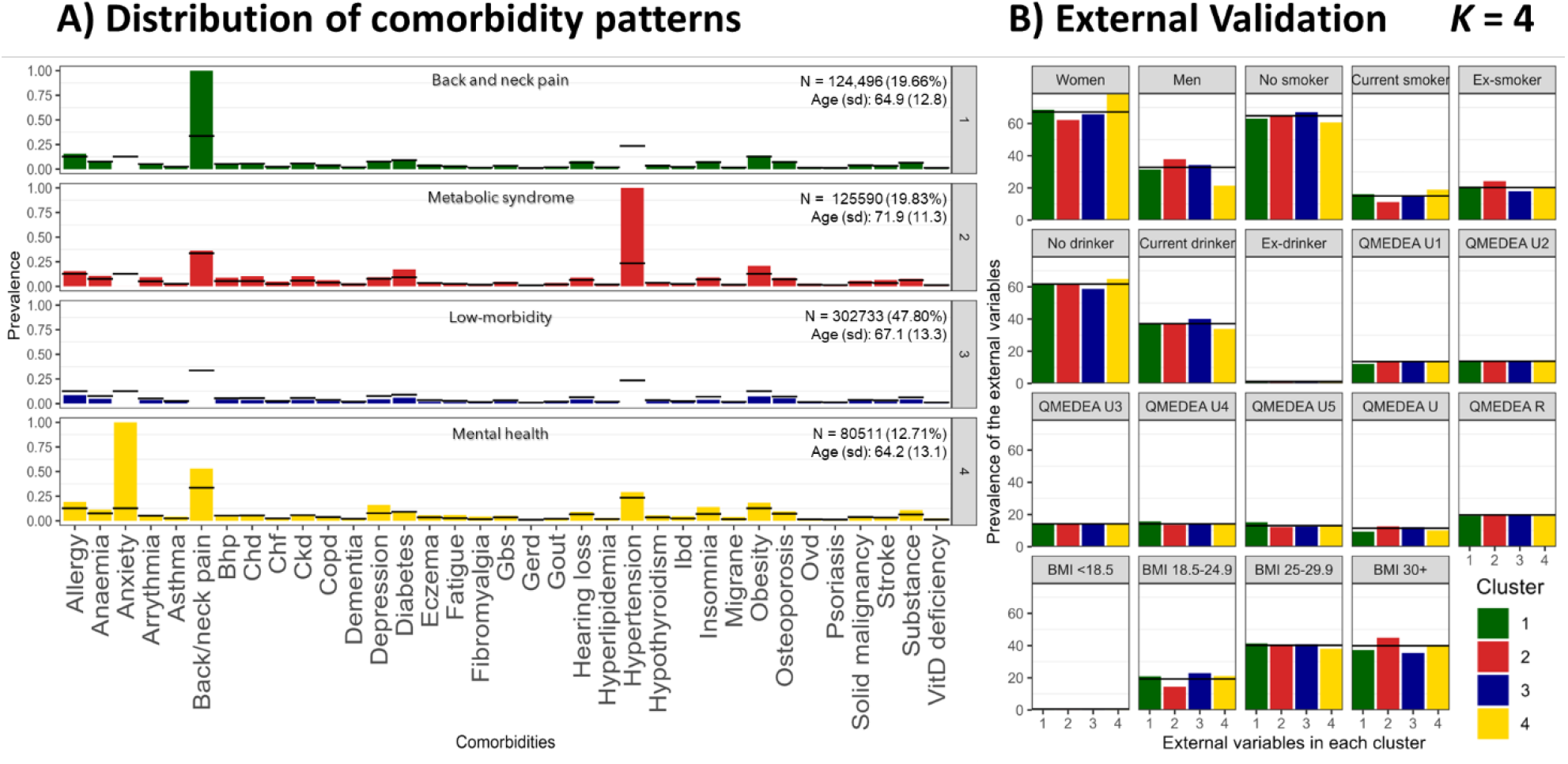
K-means cluster solution 4. A) Distribution of comorbidity patterns and B) External validation. Abbreviations: BMI, body mass index; Bhp, benign prostate hypertrophy; Chd, coronary heart disease; Ckd, chronic kidney disease; Copd, chronic obstructive pulmonary disease; Gbs, gall bladder stone; Gerd, gastroesophageal reflux disease; Ibd, inflammatory bowel disease; Ovd, other vessel diseases; Substance, substance abuse; QMEDEA, deprivation quintile index MEDEA where U is urban area (U1 is the less deprived and U5 the most), and R is rural area.

Those labelled as ‘low-morbidity’ were defined as individuals with a lower prevalence of other comorbidities compared to the general OA population. In contrast, the term ‘multimorbidity’ refers to clusters of individuals with a higher prevalence of all the listed comorbidities compared to the general OA population. Cluster of ‘metabolic syndrome’ was characterized by the presence of hypertension in all individuals, plus above average prevalence of obesity and diabetes. This group presented a higher ratio of males (37.80%) and obese individuals (44.9% had BMI ≥30) (Figure 1B). ‘Back and neck pain’ cluster was defined by the 100% prevalence of this condition in all the cluster members. Whilst ‘mental health’ label was assigned by the significant proportion of anxiety and depression: notably, all participants with anxiety were classified into this cluster. In addition, the ‘mental health’ group had the highest ratio of females (78.60%).

Supplementary Figures S2 and S3 displays the 5- and 6-cluster solutions, respectively.

### Clustering by LCA

After clustering by LCA, internal validation (Supplementary Figure S1B) using ABIC, BIC, CAIC and the likelihood ratio did not show a statistically optimal model. However, the decline ratio of the different parameters allowed us to exclude the clusters solutions equal or higher than k=6, since those did not improve model fit substantively. Evaluation of the mean posterior probability values show better discrimination for 4-cluster than 5-cluster models (Supplementary Tables S1). Hence, we selected the 4-cluster solution as tour preferred model.

When k=4, we identified the following clusters: ‘back and neck pain plus mental health’, ‘multimorbidity’, ‘low-morbidity’ and ‘metabolic syndrome’. Again, ‘low-morbidity’ refers to individuals with a lower prevalence of other comorbidities and ‘multimorbidity’ refers to individuals with a higher prevalence of all the listed comorbidities, compared to the general OA population.

The cluster with the highest proportion of participants was the ‘heathier’ (n=394,940, 62.36%), followed by ‘back and neck pain plus mental health’ (n=114,718, 18.11%), ‘metabolic syndrome’ (n=72,532, 11.45%), and ‘multimorbidity’ (n=51,140, 8.07%). Whilst our overall cohort was predominantly female (67.20%), females only made up 39.00% of the ‘metabolic syndrome’ cluster, which had the highest proportion of men. Conversely, ‘back and neck pain plus mental health’ cluster had a remarkable proportion of women (83.30%) and the youngest population (mean age [years, SD] 64.2, 12.5). In contrast, ‘multimorbidity’ cluster had the oldest population (mean age [years, SD] 79.20, 9.47). (Figure 2)

**Figure 2.**
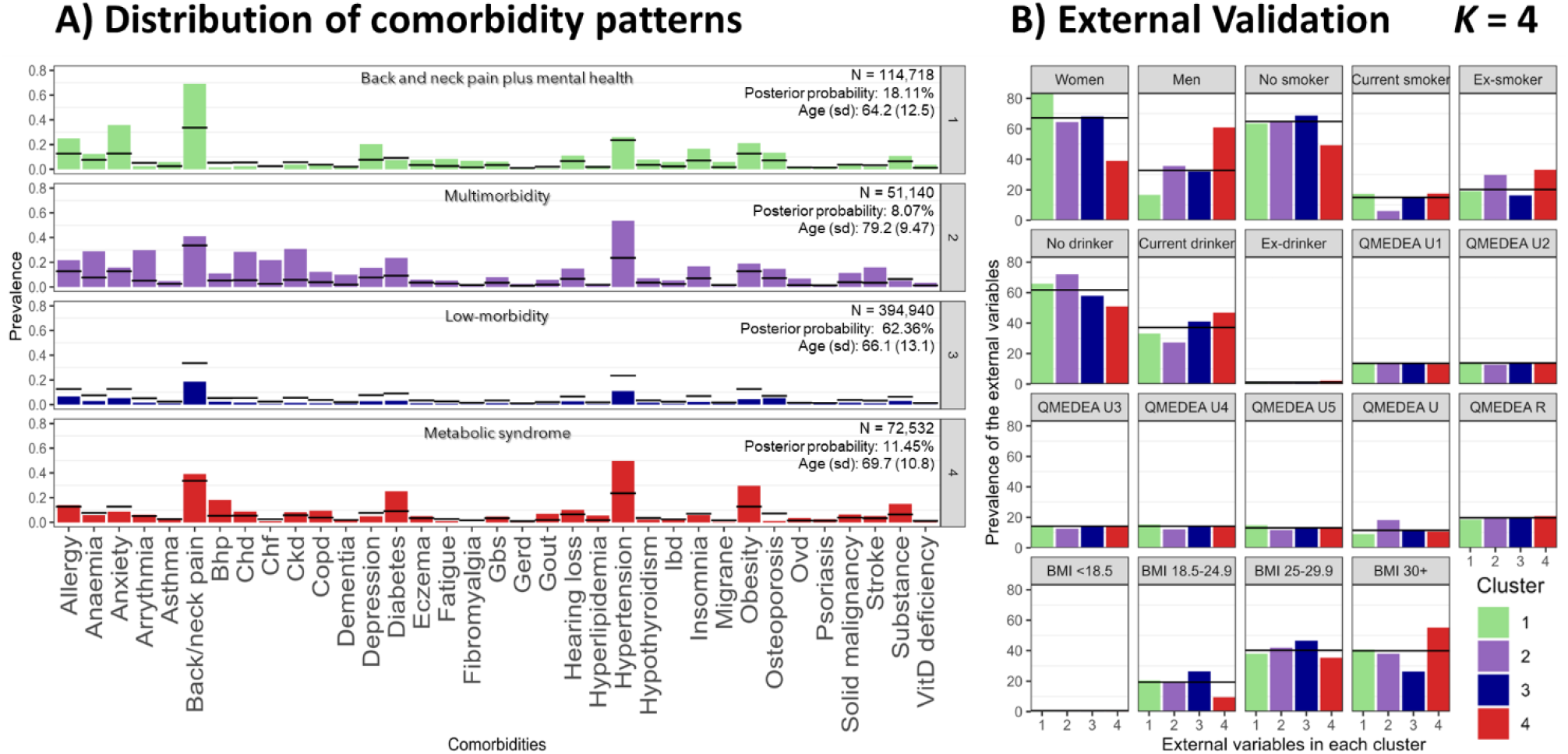
Latent Class Analysis cluster solution 4. A) Distribution of comorbidity patterns and B) External validation. Cluster colours are consistent in both sub-plots. Abbreviations: BMI, body mass index; Bhp, benign prostate hypertrophy; Chd, coronary heart disease; Ckd, chronic kidney disease; Copd, chronic obstructive pulmonary disease; Gbs, gall bladder stone; Gerd, gastroesophageal reflux disease; Ibd, inflammatory bowel disease; Ovd, other vessel diseases; Substance, substance abuse; QMEDEA, deprivation quintile index MEDEA where U is urban area (U1 is the less deprived and U5 the most), and R is rural area.

Supplementary Figures S4 and S5 reports the 5- and 6-cluster solutions, respectively.

### Survival analyses

Survival analyses for 10-year mortality (HR, 95%CI adjusted for sex and age) revealed differences between the 4-clusters identified using K-means (Table 3a) and LCA (Table 3b). The ‘low-morbidity’ cluster was used as the reference group in both analyses:

For K-means, the ‘back and neck pain’ cluster had a reduced risk of 10-year mortality (0.93, 0.91-0.95), while the ‘mental health’ (1.21, 1.18-1.24) and ‘metabolic syndrome’ (1.18, 1.16-1.20) clusters had an increased risk.

In our LCA results, all clusters, including ‘back and neck pain plus mental health’ (1.12, 95% CI 1.09-1.15), ‘metabolic syndrome’ (1.24, 95% CI 1.22-1.27) and ‘multimorbidity’ (2.19, 95% CI 2.15-2.23), had increased risk of mortality.

Supplementary Tables S2 and S3 reports the survival analysis for 5- and 6-cluster solutions in K-means and LCA, respectively.

## Discussion

Our study of 633,330 individuals with OA from the SIDIAP database is, to our knowledge, the largest to date exploring the clustering of comorbidities among individuals with a diagnosis of OA. We found that individuals with OA can be clustered based on their comorbidity patterns into groups with significantly different risks of 10-year mortality.

While we explored clustering using two separate methods, and three different cluster solutions in each of them, a number of patterns emerged: in all solutions the larger group was the ‘low-morbidity’ cluster, where patients with a new diagnosis of OA had the lowest prevalence of comorbid conditions; the ‘back and neck pain plus mental health’ groups tended to have the highest proportion of females; those designated as ‘metabolic syndrome’ groups had the highest proportion of males and the highest BMI; and the ‘multimorbidity’ groups had high mean age. Whilst age and sex varied between groups, socioeconomic status remained relatively stable. Nonetheless, the preferred solution for both clustering methods was the 4-cluster.

When K-means and LCA 4-cluster results are compared, soft classification of LCA allows higher flexibility to detect more complex patterns, such as the interaction between back and neck pain along with mental health comorbidities, or the ‘Multimorbidity’ cluster. Thus, clusters obtained by LCA better represented the behaviour and interaction within the different comorbidities (i.e., the comorbidity patterns). In addition, differences in 10-year mortality were most marked in the outgoing clusters from the LCA analyses, which may therefore be of more use when risk-stratifying patients in clinical practice.

With the caveat that more studies using different populations may shed further light on an optimal clustering solution in the future, we propose the 4-clusters identified by the LCA algorithm: ‘low-morbidity’, ‘back/neck pain plus mental health’, ‘metabolic syndrome’ and ‘multimorbidity’.

### Comparison with other literature and interpretation

A number of general patterns of multimorbidity have previously been established. Systematic reviews have identified ‘mental health’, ‘cardiovascular / metabolic’ and ‘musculoskeletal’ as common clusters of comorbid conditions^34,35^, and have found that OA with cardiovascular and/or metabolic disease is a common multimorbidity profile presenting in primary care^36^. Despite our study focussing specifically on patients with OA diagnoses, rather than the wider population, we nevertheless observed these established clusters of comorbidities in most of our analyses.

The association between cardiovascular disease and OA is established^37,38^, but whether they simply co-exist or share a common aetiology, perhaps due to age-related, inflammatory, hormonal or drug-related mechanisms, remains unclear^39^. Metabolic syndrome, classically characterised by both obesity and diabetes, is a risk factor for the development of OA through metabolic changes which affect joint function^40^. The level of obesity is also associated with the clinical severity of the disease^41^, and management guidelines therefore frequently recommend physical activity and weight loss as first-line treatment strategies in an effort to halt or slow the progression of the disease^42^. The association between musculoskeletal (especially back and neck) pain and mental health is also established^43^ and studies have shown that this link can commence early in life^44^, which may contribute to our observation that our ‘back and neck pain with mental health’ have low mean ages.

A previous study used LCA to cluster 221,807 OA patients from the UK into five groups^19^. The five groups identified were ‘low-morbidity’, ‘cardiovascular’, ‘musculoskeletal and mental health’, ‘cardiovascular and mental health’, and ‘metabolic’, which, despite differences in the specific comorbidities used for analysis, reflect our own LCA k=5 results.

Several systematic reviews have explored links between OA and mortality with varied results, likely due to underlying methodological differences between them^45-47^. In order to address some of the issues intrinsic to meta-analyses and shed further light on mortality risk in OA, a recent study used large-scale individual patient-level data from six geographically diverse cohorts and found that patients with OA-related pain, or pain and radiographic OA, had between a 35-37% increased association with reduced time to death when compared to people without OA^48^. Our data revealed that among patients with OA, their 10-year mortality risk may vary widely depending on their particular comorbidities. The largest difference seen, when compared to patients with OA who were otherwise ‘low-morbidity’, was among our ‘multimorbidity’ groups, who in some cases had almost three times the risk of 10-year mortality.

### Strengths and limitations of the study

Our study has several strengths. Firstly, we used a large established database which gathers information from >80% of its target population, allowing us to extract baseline characteristics as well as information surrounding diagnoses from a large number of participants. Secondly, our exploration of different clustering methods has allowed us to assess a variety of potential clustering results for translational potential and clinical utility. Our approach to internal and external validation, as well as assessment of mortality risk, helps to improve both the reliability and the usefulness of our findings.

Our study also has limitations. Despite the inclusion of a large number of participants, we cannot be sure that our findings are generalisable to populations in other geographic regions. Secondly, the diagnosis of OA in primary care is predominantly clinical (i.e., there is no requirement for radiographic confirmation)^42^, so there is a lack of validation of individual OA diagnoses. However, we attempted to partially mitigate this by excluding participants who had other soft tissue or bone related pathology. Furthermore, the recording of knee and hip OA within SIDIAP has previously been validated, both through comparison to self-reported physician diagnosed OA^49^, and through the analysis of free text records^50^. On the other hand, this analysis focuses on the time of OA diagnosis, so we cannot ignore the possibility that we may be observing different stages of OA, were the low-morbidity would represent an earlier stage of the diseases and the multimorbidity the other end of the spectrum. To unravel this, further work analysing patients’ trajectories is necessary.

## Conclusions

The comorbidity clusters we establish in our study for patients with a new diagnosis of OA reflect established multimorbidity patterns and are similar to those reported in a previous study using a different patient population. Such classification of patients may in the future be useful to help guide specific treatment strategies for particular groups of patients, to address both their OA as well as their other comorbidities, and may help identify ‘high-risk’ patients who require more intense input from healthcare providers. Furthermore, clustering may provide insight into shared underlying pathophysiological mechanisms between different comorbid conditions. There is a need to further validate our results in other patient cohorts, as well as research to investigate the underlying pathological mechanisms which may link the comorbidities we see in our clusters, and trials to determine the optimal treatment strategies for different groups of patients.

## Data Availability

Data that supports the findings of this study was provided by SIDIAP database. Data access is limited to researchers from public institutions, and collaboration with private organizations is only allowed for studies required by a regulatory agency or for non-commercial studies within a European project financed by the European Commission. Moreover, availability of data is subject to protocol approval by SIDIAPs Scientific Committee and Clinical Research Ethics Committee of IDIAPJGol.

## Ethics statement

In this study, all participants records were previously collected and anonymised by SIDIAP. Thus, no direct participant recruitment was done.

## Data availability statement

Data that supports the findings of this study was provided by SIDIAP database. Data access is limited to researchers from public institutions, and collaboration with private organizations is only allowed for studies required by a regulatory agency or for non-commercial studies within a European project financed by the European Commission. Moreover, availability of data is subject to protocol approval by SIDIAP’s Scientific Committee and Clinical Research Ethics Committee of IDIAPJGol.

## Acknowledgments

We thank the Patient Research Participants (PRP) members Jenny Cockshull, Stevie Vanhegan, and Irene Pitsillidou for their involvement since the beginning of the project. We would like to thank the FOREUM for financially supporting the research.

## Supplementary material

**Supplementary Figure S1.**
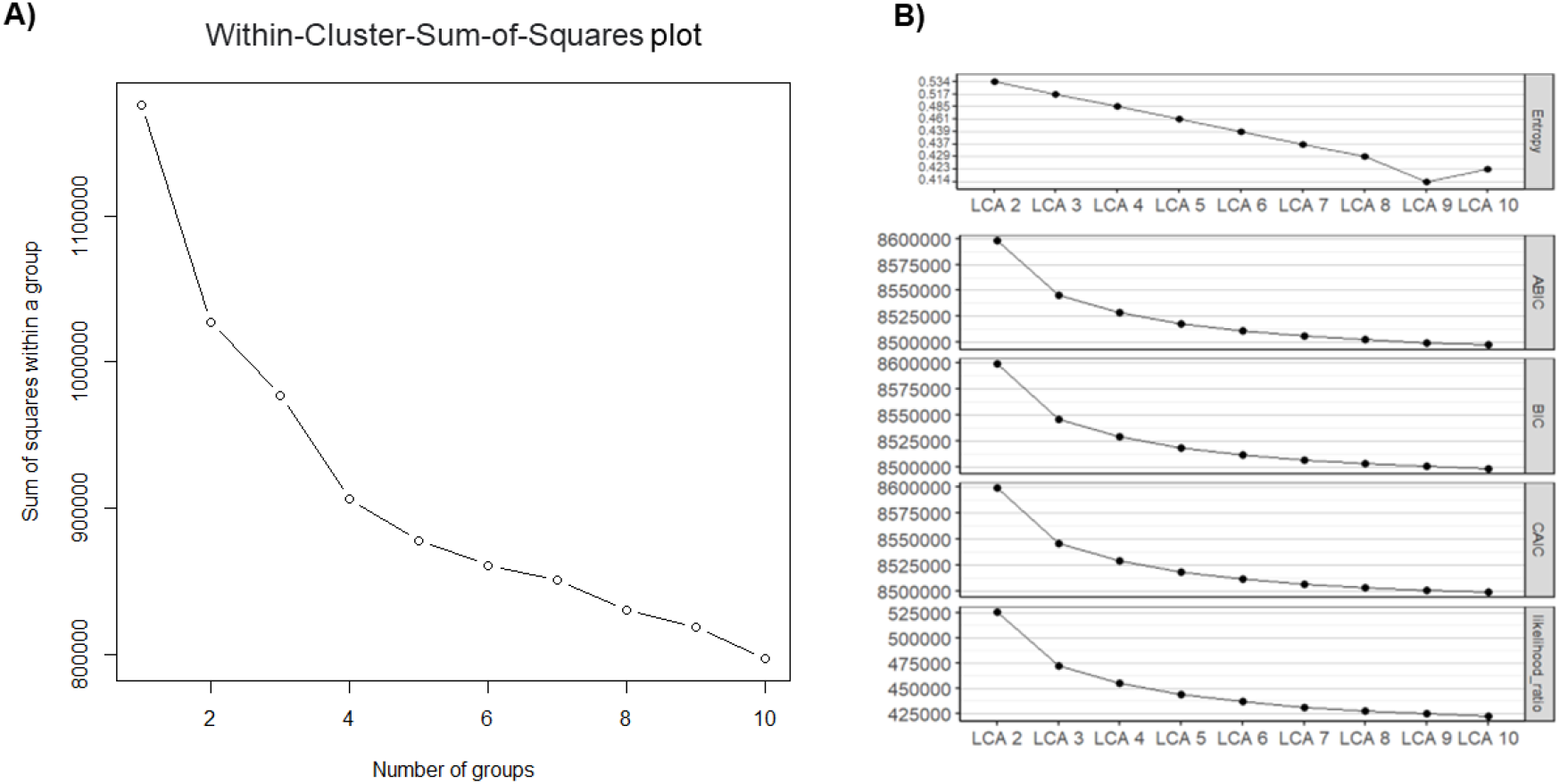
Internal validations for A) K-means and B) Latent Class Analysis solutions. The internal validation of K-means is represented by the Within-Cluster-Sum-of-Squares (WCSS) for each number of k. The internal validation of LCA is represented by the entropy R2 values, goodness of fit tests (ABIC, BIC and CAIC) and likelihood ratio for each LCA when k ranges from 2 to 10.

**Supplementary Figure S2.**
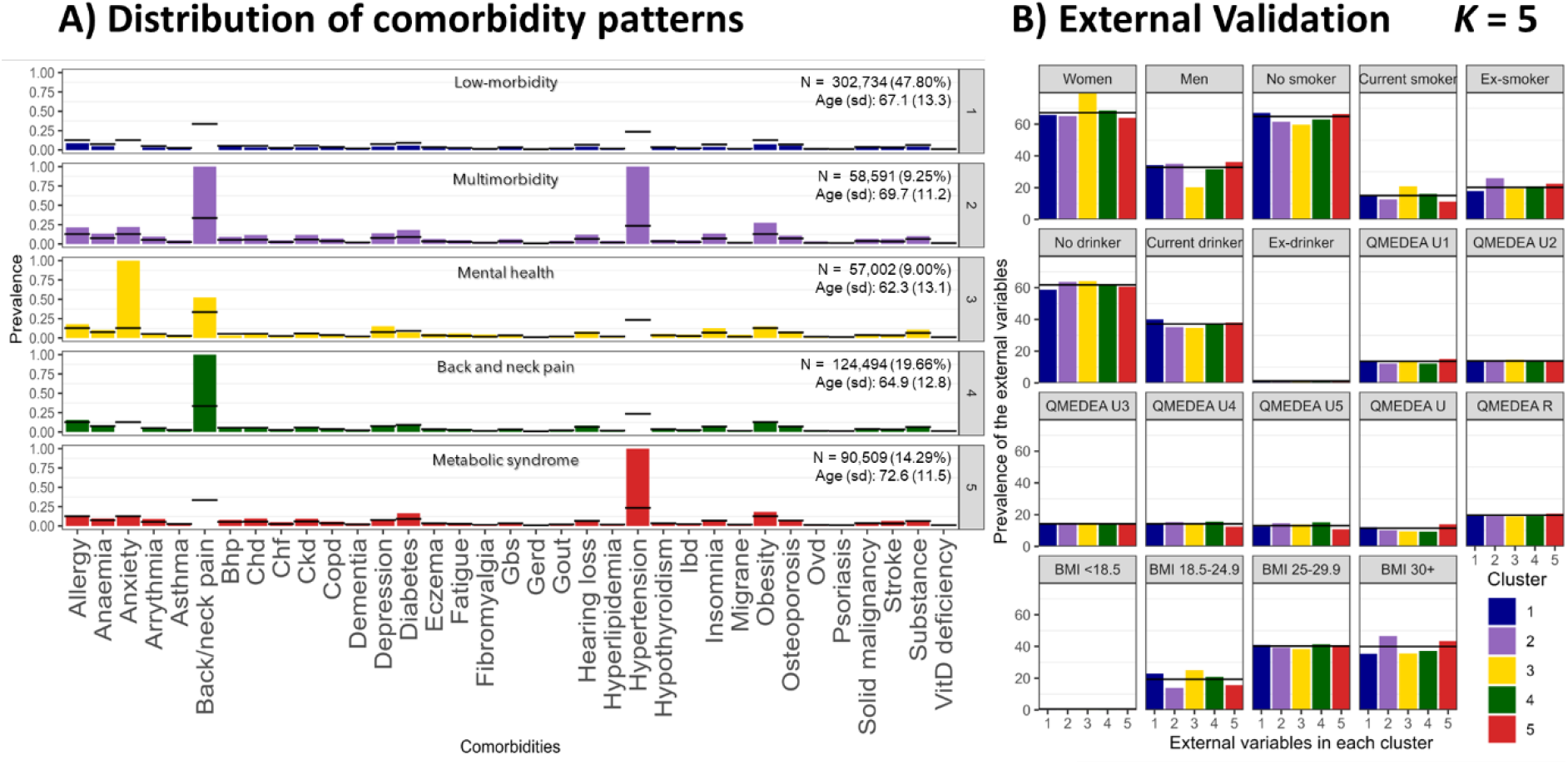
K-means cluster solution 5. A) Distribution of comorbidity patterns and B) External validation. Cluster colours are consistent in both sub-plots. Abbreviations: BMI, body mass index; Bhp, benign prostate hypertrophy; Chd, coronary heart disease; Ckd, chronic kidney disease; Copd, chronic obstructive pulmonary disease; Gbs, gall bladder stone; Gerd, gastroesophageal reflux disease; Ibd, inflammatory bowel disease; Ovd, other vessel diseases; Substance, substance abuse; QMEDEA, deprivation quintile index MEDEA where U is urban area (U1 is the less deprived and U5 the most), and R is rural area.

**Supplementary Figure S3.**
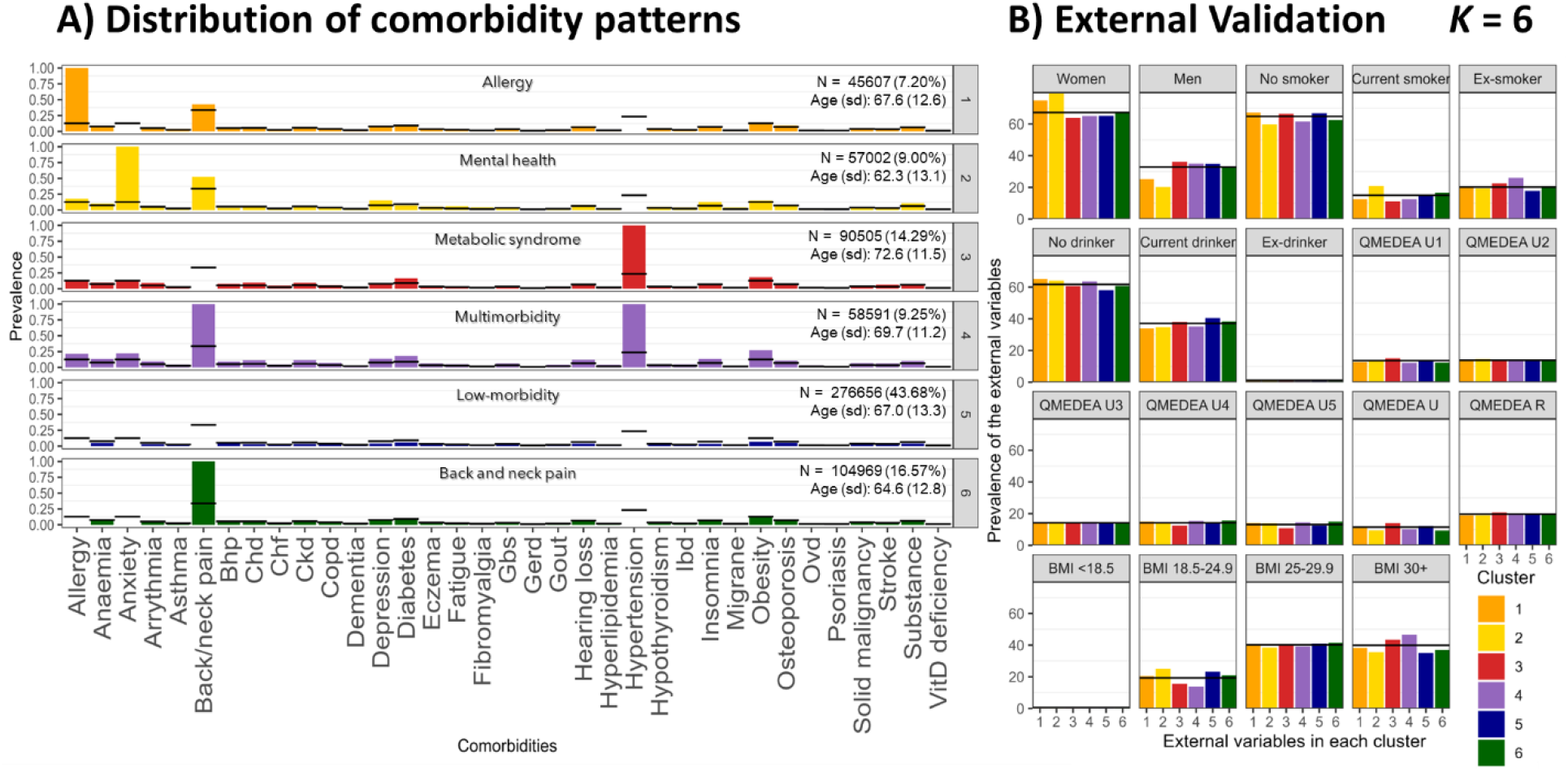
K-means cluster solution 6. A) Distribution of comorbidity patterns and B) External validation. Cluster colours are consistent in both sub-plots. Abbreviations: BMI, body mass index; Bhp, benign prostate hypertrophy; Chd, coronary heart disease; Ckd, chronic kidney disease; Copd, chronic obstructive pulmonary disease; Gbs, gall bladder stone; Gerd, gastroesophageal reflux disease; Ibd, inflammatory bowel disease; Ovd, other vessel diseases; Substance, substance abuse; QMEDEA, deprivation quintile index MEDEA where U is urban area (U1 is the less deprived and U5 the most), and R is rural area.

**Supplementary Figure S4.**
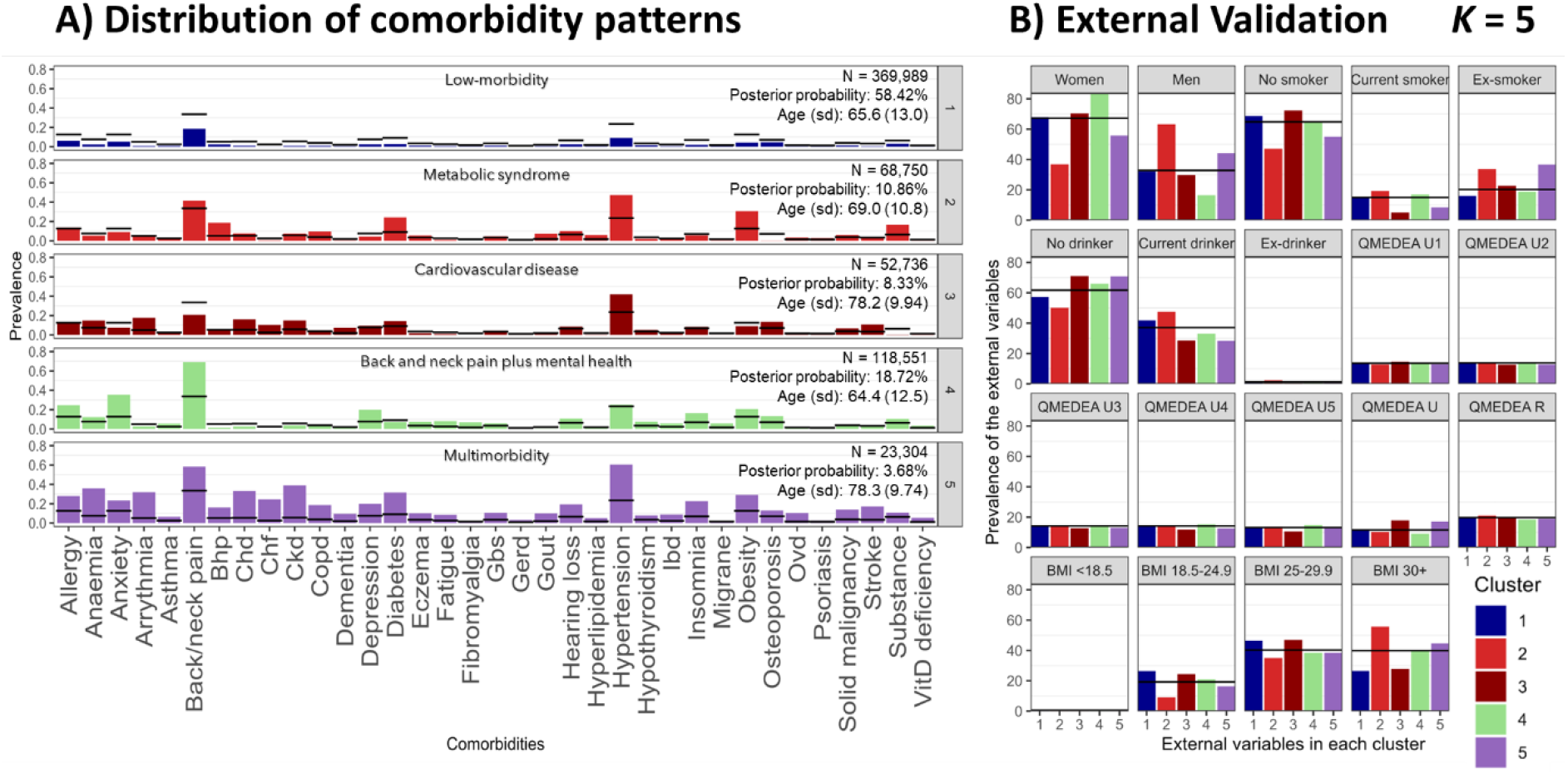
Latent Class Analysis cluster solution 5. A) Distribution of comorbidity patterns and B) External validation. Cluster colours are consistent in both sub-plots. Abbreviations: BMI, body mass index; Bhp, benign prostate hypertrophy; Chd, coronary heart disease; Ckd, chronic kidney disease; Copd, chronic obstructive pulmonary disease; Gbs, gall bladder stone; Gerd, gastroesophageal reflux disease; Ibd, inflammatory bowel disease; Ovd, other vessel diseases; Substance, substance abuse; QMEDEA, deprivation quintile index MEDEA where U is urban area (U1 is the less deprived and U5 the most), and R is rural area.

**Supplementary Figure S5.**
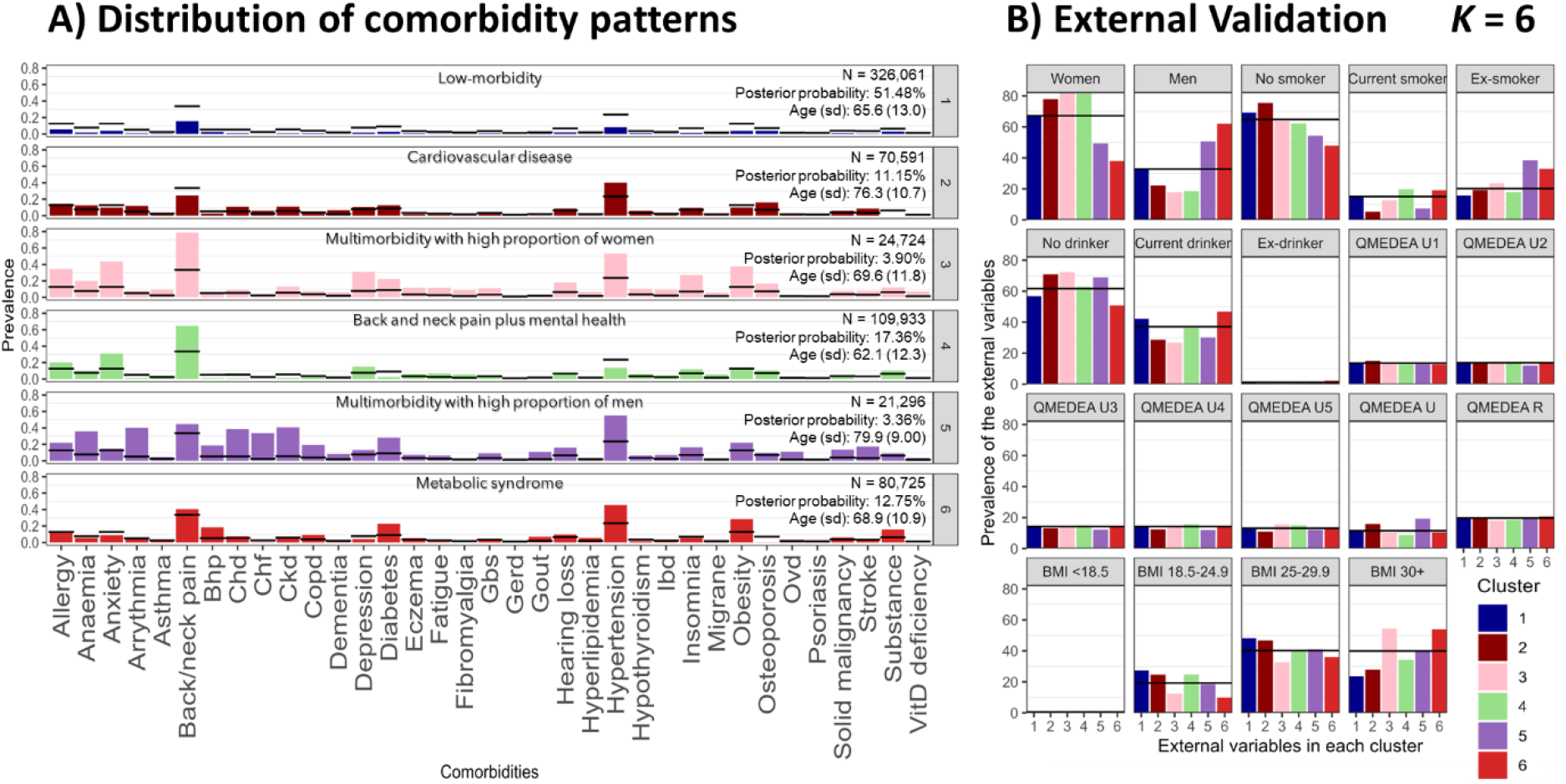
Latent Class Analysis cluster solution 6. A) Distribution of comorbidity patterns and B) External validation. Cluster colours are consistent in both sub-plots. Abbreviations: BMI, body mass index; Bhp, benign prostate hypertrophy; Chd, coronary heart disease; Ckd, chronic kidney disease; Copd, chronic obstructive pulmonary disease; Gbs, gall bladder stone; Gerd, gastroesophageal reflux disease; Ibd, inflammatory bowel disease; Ovd, other vessel diseases; Substance, substance abuse; QMEDEA, deprivation quintile index MEDEA where U is urban area (U1 is the less deprived and U5 the most), and R is rural area.

**Supplementary Table S1.**
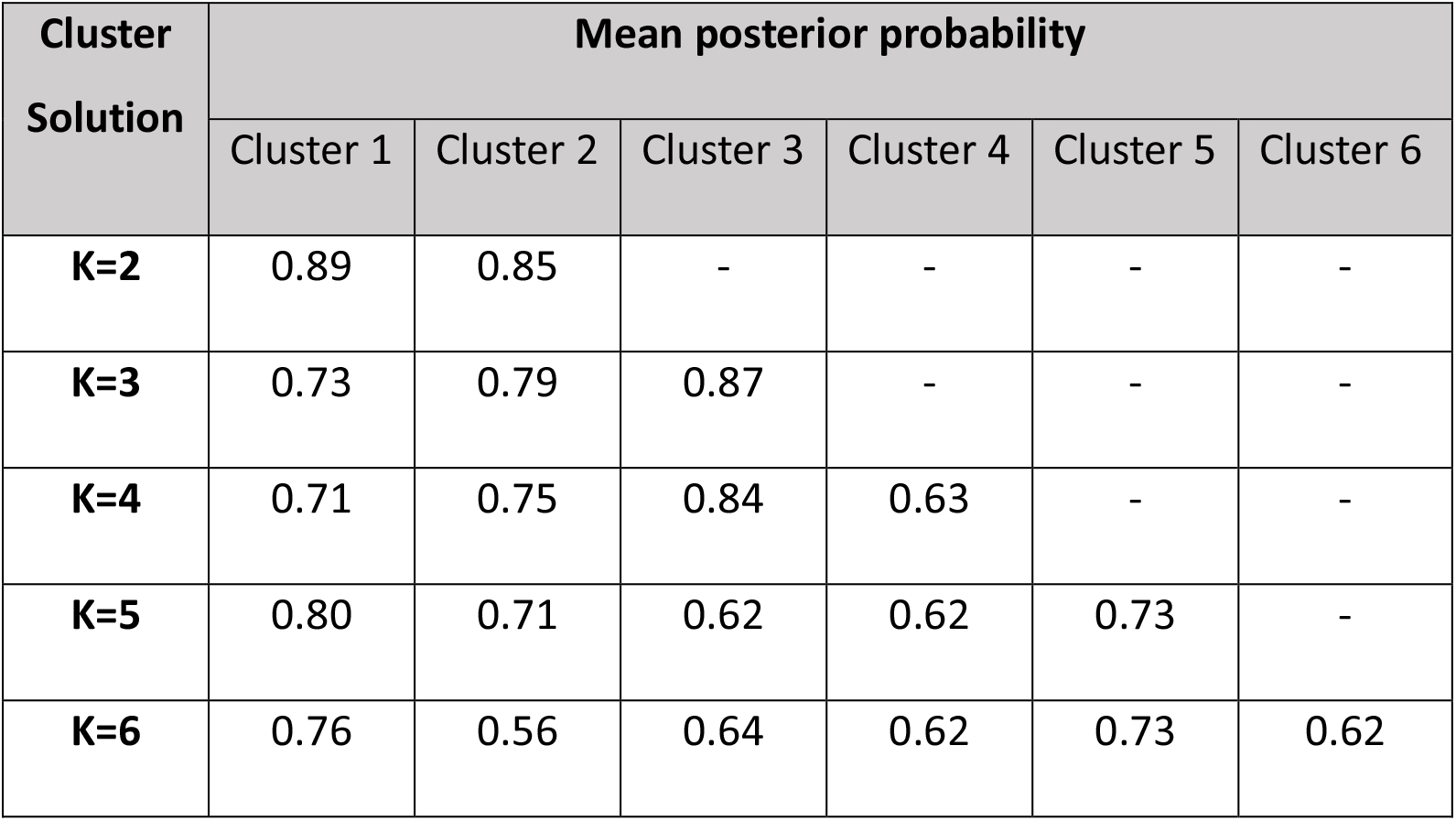
Mean posterior probability for each cluster solution in Laten Class Analysis (each person was assigned to the cluster with highest posterior probability).

**Supplementary Table S2.**
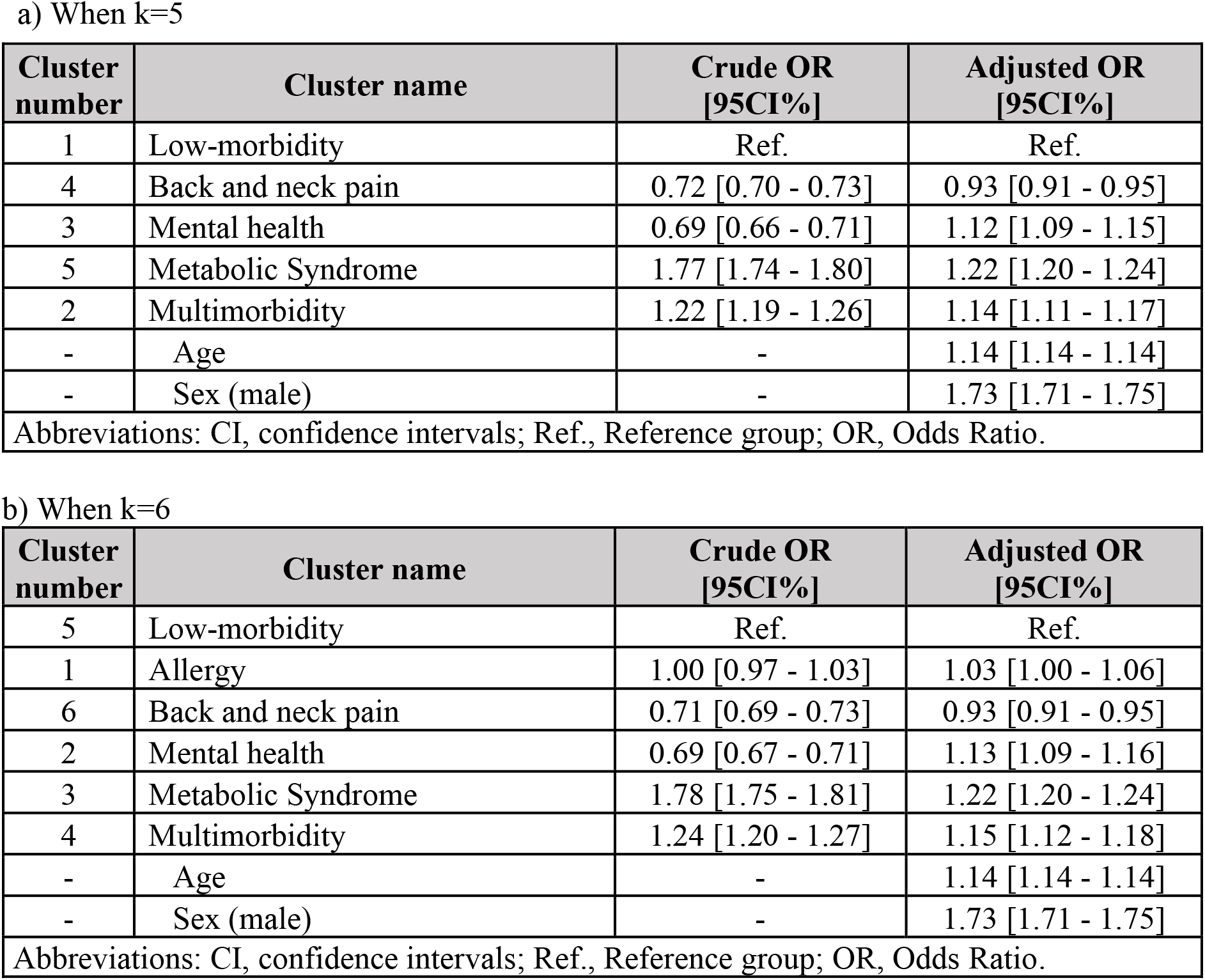
Survival analysis for 10-year mortality in a) 5- and b) 6-cluster solutions of K-means (each person was assigned to the cluster with highest posterior probability):

a) When k=5
| Cluster number | Cluster name | Crude OR [95CI%] | Adjusted OR [95CI%] |
| --- | --- | --- | --- |
| 1 | Low-morbidity | Ref. | Ref. |
| 4 | Back and neck pain | 0.72 [0.70 - 0.73] | 0.93 [0.91 - 0.95] |
| 3 | Mental health | 0.69 [0.66 - 0.71] | 1.12 [1.09 - 1.15] |
| 5 | Metabolic Syndrome | 1.77 [1.74 - 1.80] | 1.22 [1.20 - 1.24] |
| 2 | Multimorbidity | 1.22 [1.19 - 1.26] | 1.14 [1.11 - 1.17] |
| - | Age | - | 1.14 [1.14 - 1.14] |
| - | Sex (male) | - | 1.73 [1.71 - 1.75] |
| Abbreviations: CI, confidence intervals; Ref., Reference group; OR, Odds Ratio. |  |  |  |

b) When k=6
| Cluster number | Cluster name | Crude OR [95CI%] | Adjusted OR [95CI%] |
| --- | --- | --- | --- |
| 5 | Low-morbidity | Ref. | Ref. |
| 1 | Allergy | 1.00 [0.97 - 1.03] | 1.03 [1.00 - 1.06] |
| 6 | Back and neck pain | 0.71 [0.69 - 0.73] | 0.93 [0.91 - 0.95] |
| 2 | Mental health | 0.69 [0.67 - 0.71] | 1.13 [1.09 - 1.16] |
| 3 | Metabolic Syndrome | 1.78 [1.75 - 1.81] | 1.22 [1.20 - 1.24] |
| 4 | Multimorbidity | 1.24 [1.20 - 1.27] | 1.15 [1.12 - 1.18] |
| - | Age | - | 1.14 [1.14 - 1.14] |
| - | Sex (male) | - | 1.73 [1.71 - 1.75] |
| Abbreviations: CI, confidence intervals; Ref., Reference group; OR, Odds Ratio. |  |  |  |

**Supplementary Table S3.** Survival analysis for 10-year mortality in a) 5- and b) 6-cluster solutions of Latent Class Analysis (each person was assigned to the cluster with highest posterior probability):

| Cluster number | Cluster name | Crude OR [95CI%] | Adjusted OR [95CI%] |
| --- | --- | --- | --- |
| 1 | Low-morbidity | Ref. | Ref. |
| 4 | 'Back and neck pain' plus 'mental health' | 0.91 [0.89 - 0.94] | 1.15 [1.12 - 1.17] |
| 3 | Cardiovascular disease | 4.56 [4.49 - 4.64] | 1.89 [1.85 - 1.92] |
| 2 | Metabolic Syndrome | 1.69 [1.66 - 1.73] | 1.26 [1.23 - 1.29] |
| 5 | Multimorbidity | 7.03 [6.85 - 7.21] | 2.72 [2.65 - 2.79] |
| - | Age | - | 1.13 [1.13 - 1.13] |
| - | Sex (male) | - | 1.68 [1.65 - 1.70] |
| Abbreviations: CI, confidence intervals; Ref., Reference group; OR, Odds Ratio. |  |  |  |

b) When k=6
| Cluster number | Cluster name | Crude OR [95CI%] | Adjusted OR [95CI%] |
| --- | --- | --- | --- |
| 1 | Low-morbidity | Ref. | Ref. |
| 4 | 'Back and neck pain' plus 'mental health' | 0.68 [0.66 - 0.70] | 1.04 [1.01 - 1.06] |
| 2 | Cardiovascular disease | 3.46 [3.40 - 3.52] | 1.68 [1.65 - 1.71] |
| 5 | Multimorbidity with high proportion of men | 8.45 [8.25 - 8.66] | 2.84 [2.77 - 2.91] |
| 6 | Metabolic syndrome | 1.72 [1.68 - 1.75] | 1.27 [1.25 - 1.30] |
| 3 | Multimorbidity high proportion of women | 1.95 [1.86 - 2.03] | 1.62 [1.55 - 1.69] |
| - | Age | - | 1.13 [1.13 - 1.13] |
| - | Sex (male) | - | 1.68 [1.65 - 1.70] |
| Abbreviations: CI, confidence intervals; Ref., Reference group; OR, Odds Ratio. |  |  |  |

